# From Patient Voices to Policy: Data Analytics Reveals Patterns in Ontario’s Hospital Feedback

**DOI:** 10.1101/2025.01.06.25320076

**Authors:** Pourya Momtaz, Mohammad Noaeen, Konrad Samsel, Neil Seeman, Robert Cribb, Syed Ishtiaque Ahmed, Amol Verma, Dionne M. Aleman, Zahra Shakeri

## Abstract

Patient satisfaction is a central measure of high-performing healthcare systems, yet real-world evaluations at scale remain challenging. In this study, we analyzed over 120,000 de-identified patient reviews from 45 Ontario hospitals between 2015 and 2022. We applied natural language processing (NLP), including named entity recognition (NER), to extract insights on hospital wards, patient health outcomes, and medical conditions. We also examined regional demographic data to identify potential disparities emerging during the COVID-19 pandemic. Our findings show that nearly 80% of the hospitals studied had fewer than 50% positive reviews, exposing systemic gaps in meeting patient needs. In particular, negative reviews decreased during COVID-19, suggesting possible shifts in patient expectations or increased appreciation for strained healthcare workers; however, certain units, such as intensive care and cardiology, experienced fewer positive ratings, reflecting pandemic and related pressures on critical care services. ‘Anxiety’ emerged as a recurrent concern in negative reviews, pointing to the growing awareness of mental health needs. Furthermore, hospitals located in regions with higher percentages of visible minority and low-income populations initially saw higher positive review rates before COVID-19, but this trend reversed after 2020. Collectively, these results demonstrate how large-scale unstructured data can identify fundamental drivers of patient satisfaction, while underscoring the urgent need for adaptive strategies to address anxiety and combat systemic inequalities.

**Author Summary:** Understanding what patients think and feel about hospital care can lead to better health services and outcomes. We analyzed more than 120,000 patient reviews from 45 Ontario hospitals between 2015 and 2022. Our study combined natural language processing techniques to identify key concerns, including anxiety, billing difficulties, and interactions with staff. We also compared patient experiences before and during the COVID-19 pandemic, uncovering a drop in negative reviews and a rise in positive reviews, though certain units—such as intensive care—faced growing pressure. A particularly revealing finding was that hospitals located in regions with higher numbers of visible minority and low-income groups received more positive feedback before the pandemic, but this reversed after 2020. These patterns hint at deeper systemic issues, especially during times of crisis. By pinpointing the main drivers of satisfaction and dissatisfaction, our work highlights the need for healthcare services that prioritize kindness, clear communication, efficient operations, and equitable access for all. Lessons from this research could guide targeted improvements, ensuring that every patient, regardless of background or income, receives the compassionate and timely care they deserve. Our hope is that policymakers, hospital administrators, and community advocates will use these findings to shape policies that improve patient trust and well-being.

## Introduction

Patient-centered care is essential for delivering high-quality healthcare, focusing on understanding patients’ perspectives to ensure safe and effective services [1–3]. Healthcare organizations are increasingly adopting patient-centric models, recognizing that patient satisfaction directly influences treatment adherence, reduces preventable errors, lowers staff turnover, decreases hospital readmissions, and enhances overall patient engagement and health outcomes. The World Health Organization emphasizes the strong link between patient satisfaction and improved treatment adherence and health outcomes [4, 5]. However, some studies indicate that an excessive focus on patient satisfaction might compromise clinical quality by prioritizing patient preferences over evidence-based practices [6]. This conflicting evidence demonstrates the need for further research, especially in outpatient primary care where data is limited and inconsistent.

A national study of U.S. acute care hospitals found that those with high adherence to clinical guidelines also achieved better patient experience scores [7]. Interestingly, hospitals with the lowest risk-adjusted mortality rates for acute myocardial infarction had the highest patient experience scores [8]. These findings suggest that prioritizing patient experience does not necessarily undermine clinical quality. Further studies support this, showing that hospitals excelling in patient experience also report lower mortality and readmission rates and better adherence to surgical process measures [7, 9, 10]. This trend is consistent across various settings, including ambulatory care [9, 11–13].

In Canada, focusing on patient experiences and patient-centered care is critical given the country’s health system performance challenges compared to other high-income nations. A 2019 Commonwealth Fund study ranked Canada last among 11 high-income countries for timeliness and efficiency of care [14]. A tragic illustration of these systemic delays comes from a recent case where a patient in Montreal, after waiting six hours for care and ultimately leaving the emergency department, died of an aneurysm [15]. With only 127 family physicians or nurse practitioners per 100,000 Canadians as of 2023 [16], the shortage of primary healthcare providers leads to increased hospital visits during health crises and chronic condition management, potentially overwhelming the system [17]. Accessibility is further limited, with only 41% of Canadians able to see a healthcare provider the same or on next day when needed [18], and even lower rates among economically disadvantaged populations [19], pointing to the need for equitable healthcare access. These accessibility issues significantly impact patient outcomes and experiences. In the 2021-2022 fiscal year, 14.6% of mental health patients in Ontario had at least three hospital stays within a year, up from around 0.2% in 2017 [20]. This rate is 19% higher in economically disadvantaged neighborhoods, highlighting the disproportionate effects on vulnerable groups, including those with mental health conditions and lower socioeconomic status.

The COVID-19 pandemic exposed and intensified weaknesses in Canada’s healthcare system, such as provider shortages, accessibility issues, and disparities in patient outcomes [21]. Hospital overcrowding, staff shortages, and supply chain disruptions strained resources, with COVID-19 cases and deaths peaking in January 2022 [22] and totaling over 4.9 million cases and 59,000 deaths by May 2024 [23]. The pandemic showed the need for resilient healthcare systems and emphasized patient-centered care, especially for those with chronic conditions or compromised immune systems [24, 25]. Recent reports indicate there has been ‘no respite’ for Ontario patients facing long wait times and understaffed facilities, emphasizing the chronic strain on the provincial healthcare system [26]. Moreover, the mass departure of nursing professionals during the pandemic exacerbated healthcare access issues [27–29].

Research has effectively used natural language processing (NLP) and machine learning (ML) to analyze patient reviews, automating tasks such as topic classification and sentiment analysis [30–39]. However, most studies focus on online reviews on social media platforms or patient forums, lacking comprehensive analyses that incorporate equity, diversity, and inclusion (EDI) perspectives for diverse socioeconomic and demographic groups.

This study applies NLP to more than 120,000 de-identified patient reviews from 45 Ontario hospitals (2015–2022), seeking to identify factors that drive patient satisfaction, assess performance trends across units, and pinpoint care disparities for low-income and minority populations. To our knowledge, this is the first time this dataset has been analyzed using NLP and ML, providing fresh insights into patient satisfaction and equity [40]. Over 80% of hospitals had less than 50% positive reviews, revealing a decrease in positive feedback over seven years. We also found notable differences in satisfaction across regions, especially in areas with higher percentages of visible minorities and low-income communities, calling attention to the need to address inequities and establish a fair healthcare landscape.

The dataset spans January 2020 to December 2022, capturing pandemic conditions and offering a window into how public health emergencies influence patient satisfaction. Pinpointing what contributed to positive experiences during COVID-19 can help design stronger healthcare systems for future crises, ultimately promoting more responsive, inclusive, and patient-focused care in Ontario and elsewhere.

## Methods

### Dataset Description and Preparation

The US-based National Research Corporation (NRC) surveys patients in Ontario, generating a repository of self-reported hospital experiences. Through freedom-of-information requests, the Investigative Journalism Bureau (IJB) at the University of Toronto’s Dalla Lana School of Public Health acquired five years of this deidentified data, forming a collection of over 120,000 patient reviews from 45 Ontario hospitals spanning 2015 to 2022. Each record contains hospital-level data (e.g., institution name, type, units) and patient feedback (e.g., textual reviews, date of visit, sentiment rating), as well as key themes tied to satisfaction (e.g., respect, transportation, access, coordination). Multiple themes can appear in a single review [41].

Data cleaning began with tokenization and lemmatization to split text into base-form words or phrases. Non-informative tokens were removed, and spelling errors were mapped to the nearest valid word. Dates were extracted from various formats and converted to a unified JavaScript Date standard to keep consistency for web-based applications. The TextBlob library [42] then aligned sentiment valence strings and key themes to uniform spellings. Large language models (LLMs) facilitated further parsing, and the Bio-Epidemiology-NER model [43] automatically identified entities like disorders, chemicals, and drugs. We measured the distribution of hospital types, patient sentiments, extracted entities, and identified themes to uncover recurring patterns and trends. We also integrated region-based attributes for subsequent location-related analyses.

### Demographic Analysis and Hospital Categorization

We examined demographic factors using the *HealthyPlan.City* platform [44], developed by the Canadian urban environmental health research consortium (CANUE) [45, 46], which integrates environmental datasets with Census data across more than 125 Canadian cities. Using the Equity Map tool on this platform, we categorized hospitals by the proportion of visible minorities and low-income populations in adjacent regions. We set thresholds at 40% for visible minority groups and 10% for low-income communities, ensuring that roughly one-third of hospitals surpassed these cutoffs. Hospitals above these thresholds were considered more likely to serve marginalized populations, and those below were grouped separately.

To standardize inconsistent unit names across hospitals, we used a multi-step process that combined a literature review, prompt engineering, and manual checks. We first identified 27 common unit names (e.g., ‘Radiology Unit’, ‘Ophthalmology Unit’). These served as anchors for a custom prompt fed to the Claude 3 LLM [47], which mapped obvious abbreviations and handled partial matches. For instance, ‘Medical Assessment (Mail)’ was remapped to ‘Medical Assessment Unit’, and ‘H2C-Neph’ was identified as ‘Nephrology Unit’. In ambiguous cases, we consulted hospital websites for context. Unit labels like ‘G4D’ were designated as ‘Inpatient Wards’ based on the hospital’s floor-plan details. If the label remained unclear (e.g., ‘YC2BC’), the LLM proposed ‘Inpatient Wards’ as a plausible match, with a note that further details could refine the classification. This structured approach ensured coherent naming conventions across diverse hospital data.

### COVID-19 Period Temporal Segmentation

To evaluate changes in patient experiences during the COVID-19 pandemic, we labeled reviews from January 2020 to December 2022 as ‘COVID-19 period’. This window aligns with the World Health Organization’s public health emergency designation from January 30, 2020, to May 5, 2023 [48, 49], making it possible to compare pre-pandemic and pandemic data consistently. We applied this temporal division to the entire dataset, enabling clear comparisons of patient sentiment and hospital performance across different timeframes.

## Results

### Preliminary Results

Our analysis of patient reviews from 45 Ontario hospitals shows that more than 80% of these facilities received under 50% positive reviews, suggesting significant potential to enhance patient satisfaction. The total volume of reviews varied sharply from 2015 to 2022, rising from 541 reviews in 2015 to a peak of 26,243 in 2020 before dropping to 1,708 in 2022 (Fig 1). These fluctuations may reflect shifts in patient engagement through online feedback platforms, combined with an uptick in healthcare discussions during the early pandemic phase. The lower numbers for 2022 could stem from the dataset including reviews only until July of that year.

**Fig 1.**
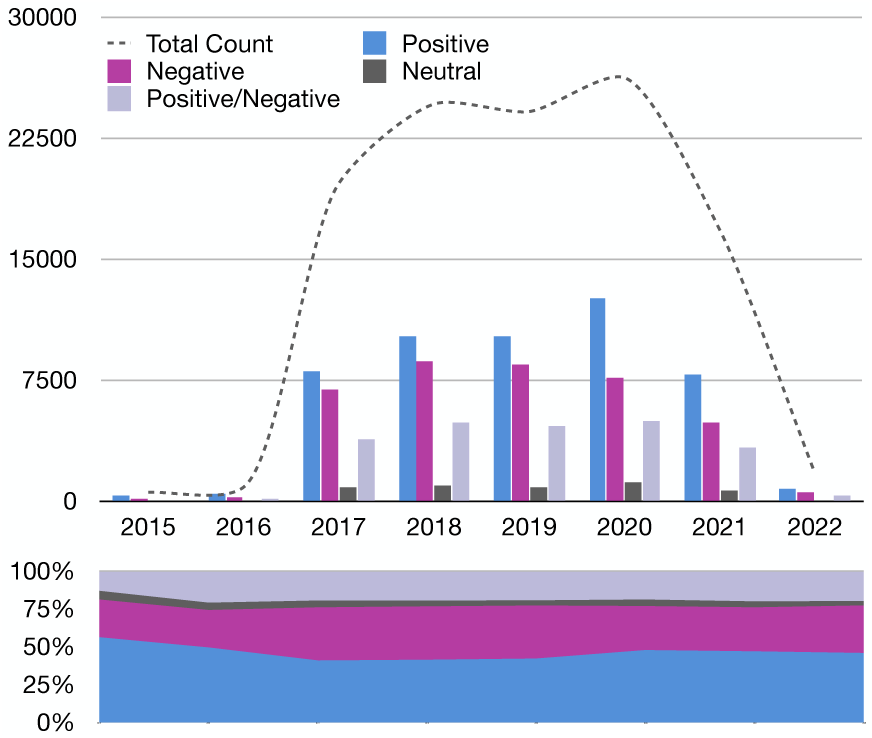
Temporal analysis of review sentiment distribution (2015-July 2022). The upper panel presents a grouped bar chart depicting the absolute frequency of positive, negative, and neutral reviews annually. The lower panel illustrates the relative proportion of each sentiment category as a stacked percentage bar chart. This dual representation reveals both absolute and relative changes in review sentiments over time, signaling trends in patient feedback and potential external influences.

Fig 1 illustrates trends in sentiments from 2015 to 2021. Positive reviews were generally the largest category, ranging from 56.19% (2015) to 40.84% (2017), while negative reviews spanned 24.77% to 35.31%. Neutral reviews stayed below 5.73%. Although positive feedback climbed from 2017 to 2020, it slipped from 47.7% in 2020 to 45.8% in 2021. Concurrently, negative feedback rose from 29.11% to 31.4%. This result points to the pandemic’s probable impact on patient satisfaction. Restricted visitation, overworked staff, and increased safety precautions may have decreased positive experiences while fueling more negative reactions, which aligns with similar findings reported during the pandemic in other healthcare systems [48, 49].

Word-frequency analysis suggests that staff behavior, rather than clinical outcomes, drives both favorable and unfavorable reviews. Terms such as ‘nurse’ and ‘staff’ were widespread. Positive responses frequently included words like ‘professional’, ‘helpful’, and ‘caring’, indicating that qualities like competence and empathy form a foundation for trust. Negative reviews often cited ‘waiting’, ‘time’, ‘night’, and ‘hour’, suggesting long waits and logistical delays are a major source of frustration. An elevated mention of ‘doctor’ implies an opportunity to improve interactions with physicians, which can shape overall care perceptions. These insights signal that interpersonal communication and efficient operations are top priorities for boosting satisfaction, mirroring global perspectives on patient-centered care [11].

### Trends in Negative and Positive Reviews

Analysis of patient reviews across different hospital units shows varied patterns in satisfaction. Some units reported more negative sentiments, while others held steady or improved positive ratings. The percentages listed below reflect the share of each unit in the total volume of positive or negative feedback.

The emergency unit reported a rise in negative reviews, moving from 36.1% in 2017 to 44.3% in 2021. This aligns with recent data showing Ontario’s ‘hallway health care’ challenges are worse than ever, as crowded hallways and bed shortages persist [50]. Longer wait times, crowded facilities, and understaffing might explain this spike. Reports from Ontario’s Ministry of Health have noted that many emergency departments face staff shortages, which can amplify patient dissatisfaction. Meanwhile, the maternity and pediatric units maintained low negative-review percentages. The maternity unit’s share of negative reviews shrank from 1.8% in 2017 to 0.3% in 2021, and the pediatric unit’s share went down from 4.1% to 1.1%. The medical/surgical unit also showed consistently lower negative reviews, varying from 4.1% in 2017 to 2.5% in 2021. This stability might reflect readiness for pandemic-related demands. Some local reports attribute this resilience to well-defined protocols that helped staff respond to workload surges without compromising care quality.

Although the emergency unit struggled with increased negative perceptions, it also showed a rise in positive reviews, climbing from 30.6% in 2017 to 44.7% in 2021. This shift may point to effective problem-solving measures in emergency care. In contrast, the maternity unit faced a drop in positive reviews, going from 3.6% in 2017 to 0.9% in 2021, while the pediatric unit’s positive reviews dropped from 5.8% to 2.5%. COVID-19 restrictions, such as limited visitors and face coverings, may have affected maternity and pediatric feedback [51–53]. These findings suggest a complex balance between providing safe care and addressing patient preferences, especially in units focused on women’s and children’s health.

### Key Clinical Entities in Patient Reviews

Named entity recognition (NER) uncovered trends in frequently mentioned clinical topics in negative reviews. ‘Pain’ stood out as the most frequent term (Fig 2(a)), totaling 4,905 mentions (Fig 2(b)). Patients are often asked to describe their pain, which may drive these high counts.

**Fig 2.**
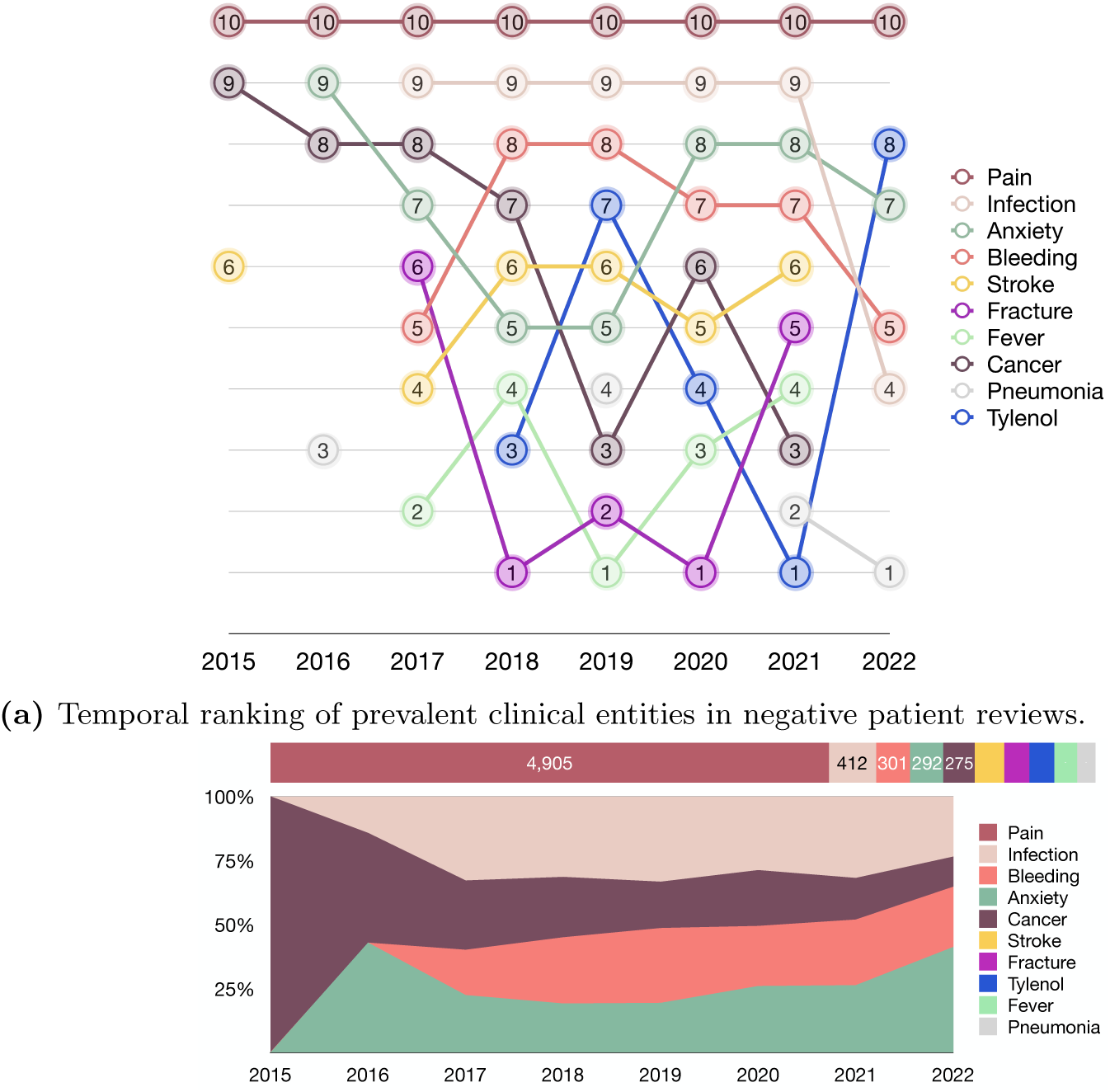
Common entities in negative patient reviews (2015-July 2022). (a) Bump chart showing annual ranking trends of frequently mentioned clinical entities, with 10 representing the most frequent. (b) Stacked bar chart (top) depicting raw counts and normalized area chart (bottom) showing the relative prevalence of Infection, Bleeding, Anxiety, and Cancer, illustrating absolute frequencies and proportional trends.

’Infection’ was second most mentioned from 2017 to 2020 before dropping to seventh in 2022, indicating changing patient concerns over time. ‘Anxiety’ rose from being unranked in 2015 to among the top five from 2020 onward, reflecting a sharper focus on mental health in hospital reviews during the pandemic. ‘Bleeding’ and ‘stroke’ remained consistent, while ‘cancer’ and ‘fracture’ shifted in importance. These changes may match evolving hospital services or patient demographics. They also emphasize the need for flexible care models that respond to new demands, such as rising orthopedic cases or advanced cancer treatments.

In negative reviews, leading concerns included ‘pain’, ‘infection’, ‘bleeding’, ‘anxiety’, and ‘stroke’. In positive reviews, ‘pain’, ‘cancer’, ‘stroke’, ‘anxiety’, and ‘infection’ took the top spots. The changing positions of these terms between the pre-COVID and COVID-19 periods (Fig 2(b)) show that patient priorities shift under different conditions. Word frequency analysis also reveals that beyond clinical terms, patient satisfaction is linked to staff interactions and efficient service. Positive reviews feature words such as ‘professional’ and ‘caring’, whereas negative reviews include ‘waiting’ and ‘time’. This points to the importance of addressing operational hurdles and ensuring compassionate communication to strengthen patient trust.

### Performance During COVID-19

Our analysis found that the percentage of negative reviews during the COVID-19 period was 6% lower than during the non-COVID period, while the percentage of positive reviews was higher. This pattern may reflect greater public sympathy for hospitals amid extreme pandemic conditions [54], or effective efforts to adapt services to evolving challenges.

The analysis of key themes showed that during the COVID-19 period, ‘positive recognition’ had the highest percentage of positive reviews (73.18%), while ‘billing/accounting’ had the highest percentage of negative reviews (63.84%). In the non-COVID period, ‘positive recognition’ also had the highest percentage of positive reviews (73.77%), and ‘billing/accounting’ had the highest percentage of negative reviews (68.34%). This result implies that while many patients appreciated staff acknowledgment, billing issues remained a major source of dissatisfaction.

Comparisons of positive review percentages (Figure 3) suggest that ‘ICU/CCU’, ‘cardiology’, ‘radiology’, ‘families/friends’, ‘social services’, ‘discharge’, ‘information/education’, and ‘positive recognition’ received fewer positive reviews during COVID-19, suggesting the pandemic’s uneven impact. ICU/CCU and cardiology departments, for instance, were pushed to the limit by surging critical cases, possibly dampening patient satisfaction. Other themes showed higher positive review rates, pointing to the adaptability of some services.

**Fig 3.**
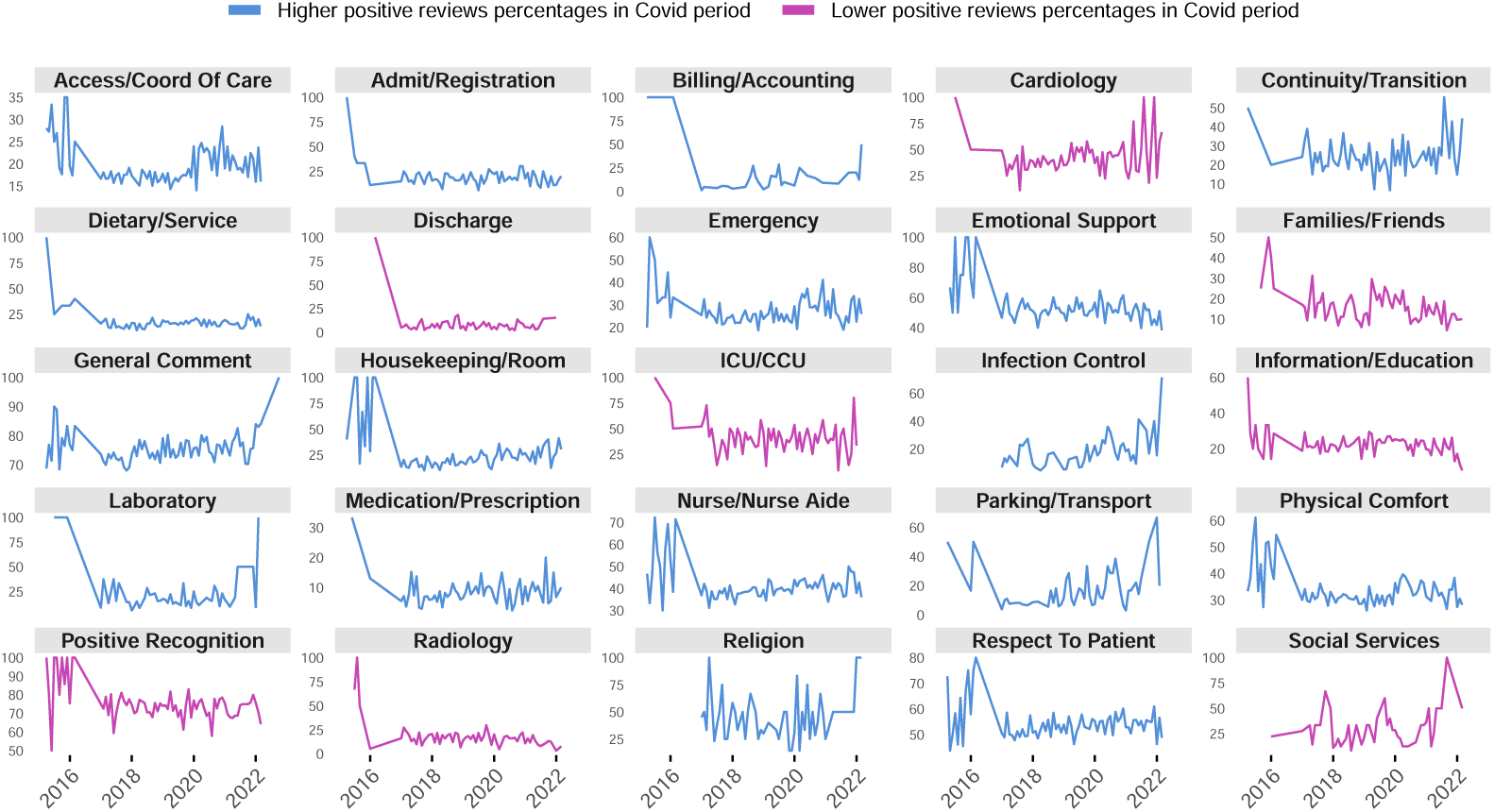
Positive review percentages for key hospital themes before and during COVID-19. Themes like ‘ICU/CCU’, ‘cardiology’, and ‘radiology’ experienced declines, indicating significant challenges during the pandemic. In contrast, other themes showed increased positive reviews, reflecting hospitals’ resilience in enhancing patient experiences amid unprecedented challenges.

The Comet chart of negative review changes (Figure 4) showed that pediatric units had more negative reviews tied to ‘families/friends’, suggesting complications around family-centered care. In inpatient units, ‘social services’ and ‘parking/transport’ had higher negative review counts, implying that support services and accessibility remained problematic. ‘Infection control’ was less common in negative reviews during COVID-19, hinting that hospitals improved or clarified safety protocols. These findings emphasize the value of consistent family support, seamless accessibility, and strong support services, while preserving infection control.

**Fig 4.**
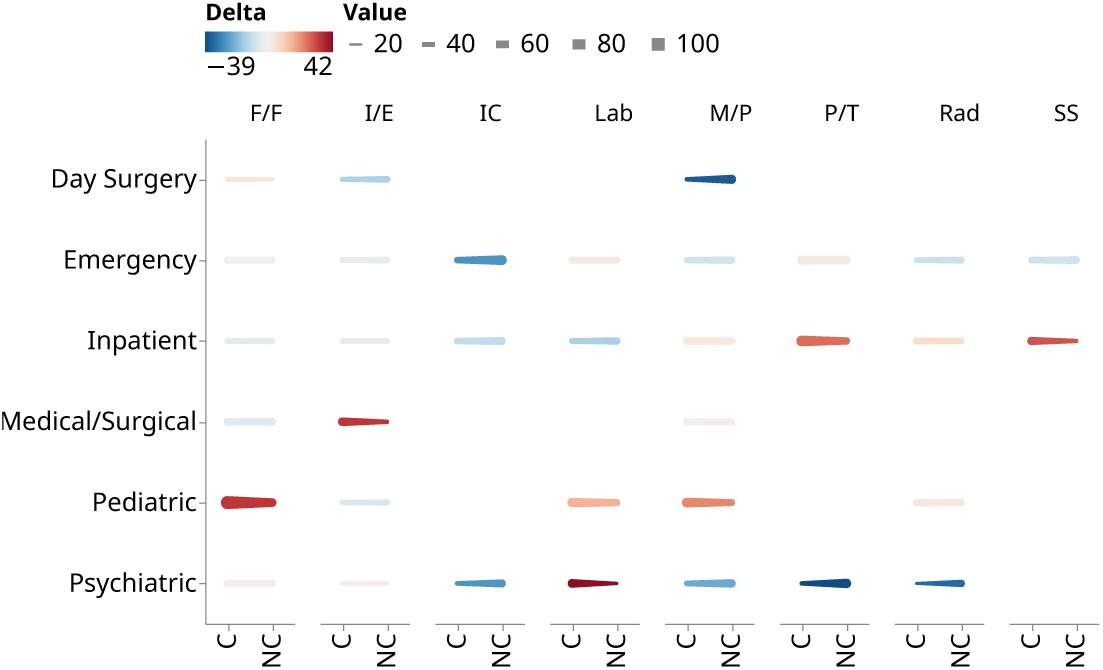
Changes in positive review percentages for key themes in different hospital units during COVID-19 and non-COVID periods. Key themes are abbreviated as follows: information/education (I/E), medication/prescription (M/P), infection prevention & control (IC), families/friends (F/F), parking/transport (P/T), social services (SS), laboratory (Lab), and radiology (Rad). ‘C’ and ‘NC’ on the x-axis represent COVID-19 and non-COVID periods, respectively.

Positive review percentages rose for most hospital units during COVID-19 compared to the non-COVID period, with dentistry as a notable exception (31.25% vs. 20.57%). Cardiology and day surgery units saw relatively high positive ratings during COVID-19 (51.47% and 56.01%, respectively) (Figure 5 (a)), which may reflect stronger infection control measures and patient safety protocols [55]. For dentistry and radiology, lower positive reviews (and higher negative ones) may stem from restricted service availability or heightened anxiety about close-contact procedures.

**Fig 5.**
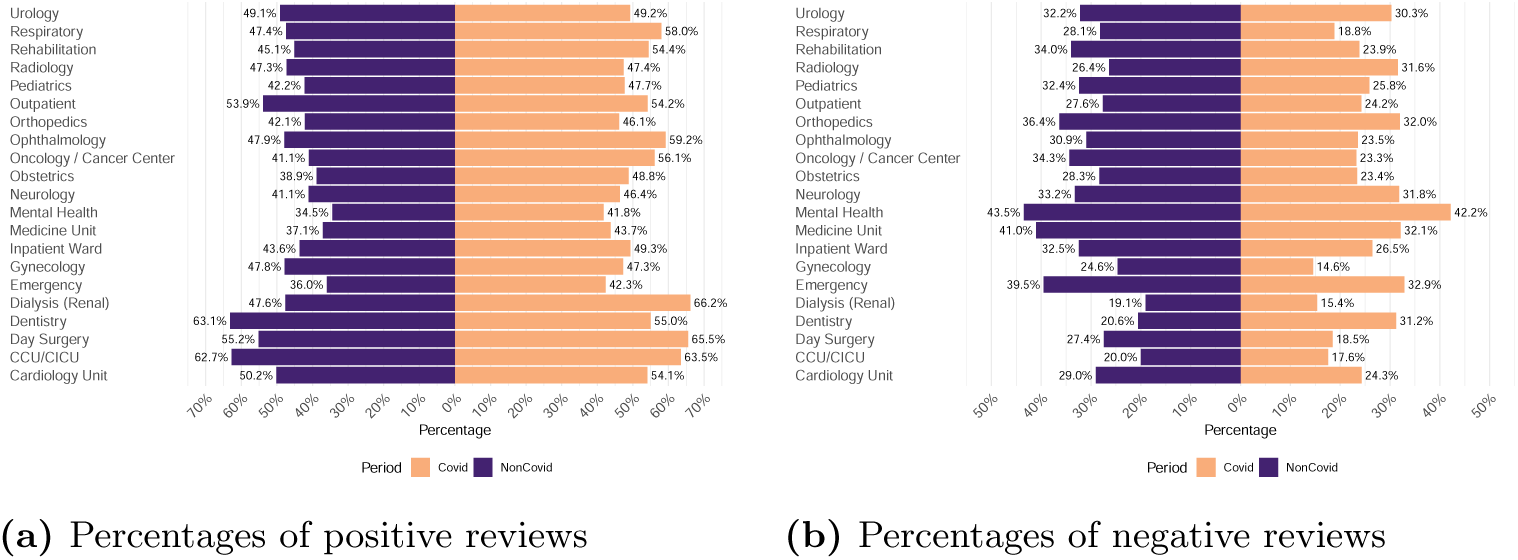
Positive and negative review percentages across different hospital units (COVID-19 vs. non-COVID Periods). This dual representation captures the shifts in patient satisfaction and dissatisfaction across different hospital units due to the pandemic, revealing critical insights into the impacts on healthcare services and areas needing targeted improvements.

Emergency and inpatient units had lower negative percentages during COVID-19 (32.81% and 26.89%) than before (38.91% and 31.92%), hinting that fewer non-urgent visits or increased public empathy may have contributed to slightly better perceptions. These results illustrate how crisis responses can shape patient satisfaction [56]. Dentistry and radiology, on the other hand, indicate unmet needs that demand targeted solutions. A recent multicenter survey reported similar diverging trends, reinforcing the idea that some units handled acute pressure better than others [40].

### Equity, Diversity, and Inclusion (EDI) Analysis

Figure 6 shows the broader demographic and sentiment patterns across six Toronto regions. Visible minority percentages range from *<*20% to *>*90%, yet high negative review rates do not always align with areas of higher minority density, indicating that other elements like socioeconomic conditions or local health resources may be pivotal. Notably, non-COVID care consistently received more negative reviews than COVID-focused care, suggesting persistent obstacles in everyday healthcare. Positive review rates vary by region, which can reflect possible differences in staffing or local health policies.

**Fig 6.**
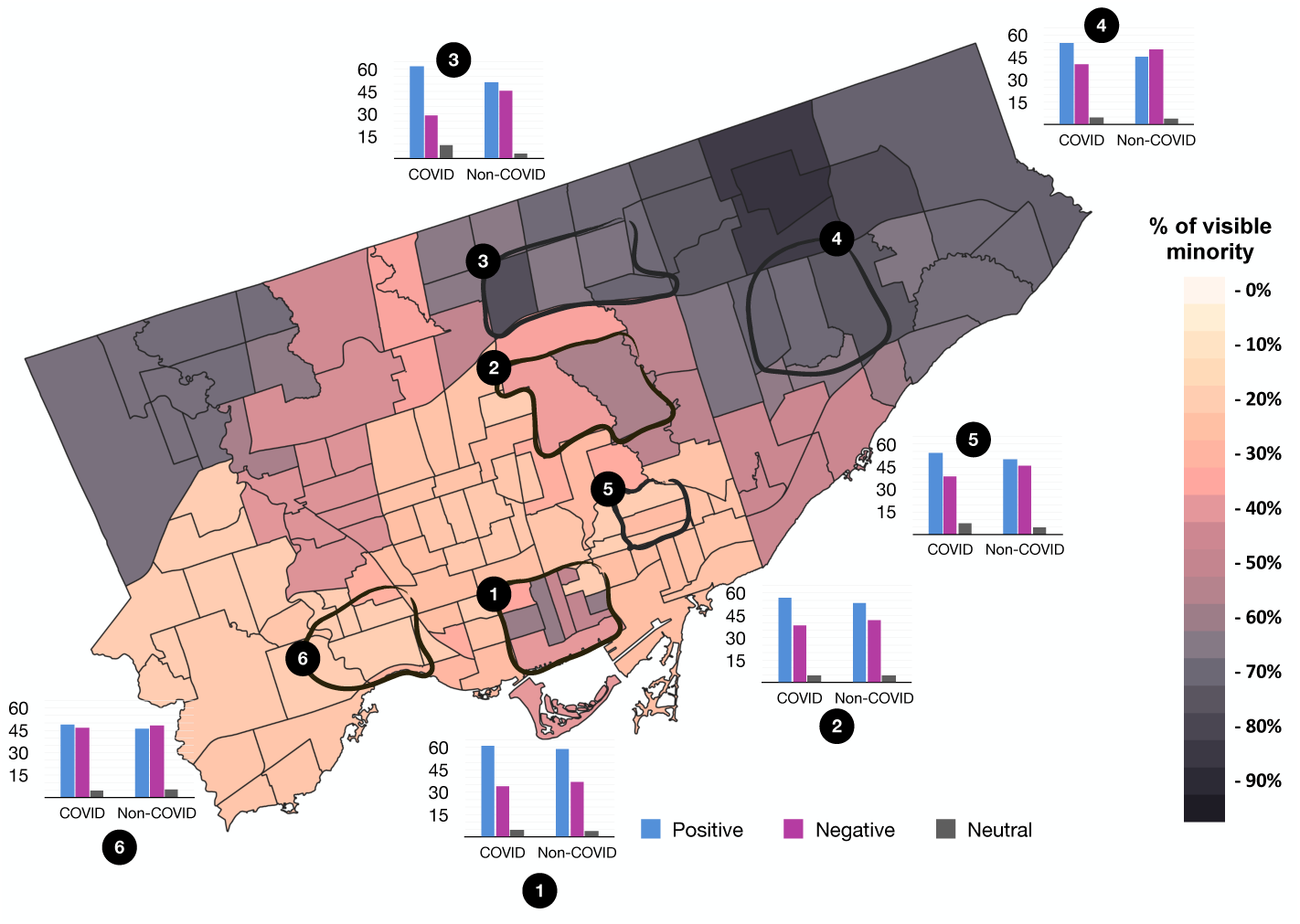
Visible minority demographics and hospital review sentiments in Toronto. A heatmap of visible minority percentages across Toronto’s six hospital regions, alongside bar charts depicting patient review sentiments. Regions 1-4 exhibit *>*40% visible minority populations, while regions 5-6 show *<*40%.

### Visible Minority Analysis: Pandemic Exacerbates Existing Inequalities

From 2015 to 2020, hospitals in high-minority areas generally performed better. During 2021–2022, this trend reversed, with these hospitals showing 44.47% positive reviews in 2021 (vs. 48.01% in low-minority areas) and 41.94% in 2022 (vs. 47.44% in low-minority areas). Negative reviews ticked upward for high-minority regions (31.32% in 2021 vs. 28.04% in low-minority regions). The gap persisted into 2022 but with a slight dip in high-minority negative reviews. This shift may reflect heavier burdens on high-minority areas, coupled with existing disparities and changing patient expectations.

### Low-Income Analysis: Economic Vulnerability Amplifies Healthcare Disparities

Hospitals in higher low-income areas had better reviews from 2015 to 2020 but faced a sharp downturn in 2021–2022. Positive reviews dropped from 44.74% to 40.95%, while negative reviews rose from 30.46% to 31.61% (Figure 7 (b)). Chronic underfunding, limited crisis response capacity, and higher COVID-19 exposure risks for economically vulnerable groups may fuel these shifts. Strengthening community partnerships and investing in better access to care could alleviate these pressures.

**Fig 7.**
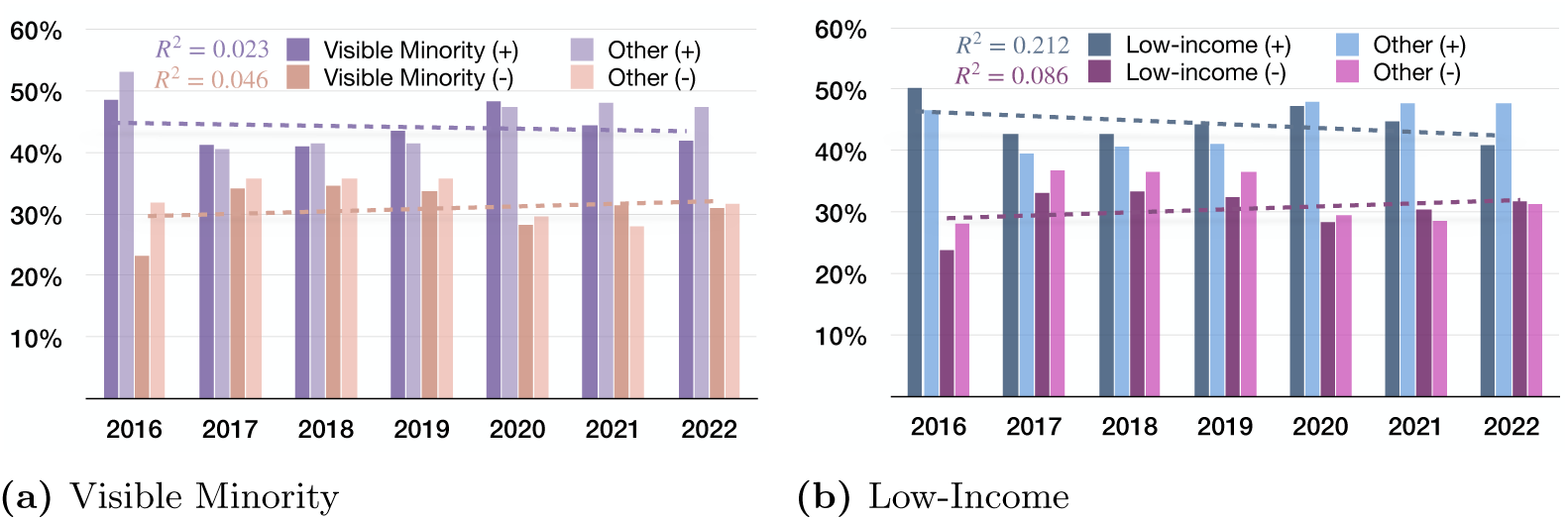
Temporal trends in patient feedback across demographic groups (2016-July 2022). Percentage of positive and negative comments from (a) visible minority groups and other demographic categories and (b) low-income and other income categories. Dashed lines represent linear regression fits with *R*^2^ values indicating model fit quality.

## Discussion

### Key Observations

Below we present the main findings from our analysis, drawing attention to patterns that could reshape healthcare practices and policies.

### Patient Satisfaction as a Launchpad for Change

In today’s digital age, over 70% of patients research hospital reviews before deciding where to receive care, a significant increase from just 25% in 2013 [57], demonstrating the growing impact of online feedback on patient decisions. However, despite this trend, a recent report reveals that Ontario hospitals, while collecting substantial patient feedback, kept the findings confidential, leaving patient pleas for improvements unheard [40]. The finding that over 80% of hospitals had less than 50% positive reviews signals a pressing need for major overhaul in our healthcare system, aligning with Canada’s last-place ranking among 11 high-income nations in timeliness and efficiency of care [58], indicating a critical need for broad reform. The COVID-19 pandemic made these issues worse, eroding a once-improving patient satisfaction trend. From 2020 onwards, the gap between negative and positive reviews narrowed, suggesting a growing sense of dissatisfaction. This decline stems from intense pressure on healthcare resources, rapid adoption of new safety measures, and rising patient anxiety during an unpredictable global crisis [21]. Nevertheless, the high proportion of negative reviews may partly reflect self-selection bias, where patients who experience dissatisfaction are more inclined to leave feedback. Furthermore, the data collection methods used by hospitals or external review platforms could amplify this effect. However, the consistent pattern of concerns across multiple hospitals suggests that significant systemic issues remain unaddressed, and the urgency of such issues is not diminished by potential sampling biases.

### Impact of Staff Behaviour on Patient Perceptions

Our analysis shows that human interaction is central in healthcare. Positive reviews often mentioned staff attributes such as ‘professionalism’, ‘helpfulness’, and ‘attentiveness’, aligning with research that calls attention to communication, empathy, and respectful treatment as key factors in patient experiences and outcomes. Negative reviews, in contrast, frequently pointed out long wait times, which is consistent with studies showing that excessive waits can lower satisfaction [59, 60].

These patterns underline the importance of well-rounded training initiatives that target clinical skills, interpersonal engagement, emotional intelligence, and patient-focused dialogue. Creating a culture of compassion and respect can boost patient satisfaction and overall well-being [61–63]. The frequent mention of ‘doctor’ in negative reviews is especially concerning, suggesting issues such as high workloads, time constraints, or insufficient interpersonal training. Further investigation is warranted, possibly through targeted surveys, focus groups, and robust feedback analysis. We advocate for holistic training, more inclusive workplace cultures, effective strategies to shorten wait times [64, 65], and real-time monitoring of patient feedback to address these critical issues.

### Challenges and Adaptations During the COVID-19 Pandemic

The COVID-19 pandemic introduced extraordinary hurdles for healthcare systems. Our analysis showed evidence of changes in patient satisfaction trends during this period. Negative reviews often included ‘anxiety’ and ‘infection’, reflecting the pandemic’s mental and physical toll on patients [21]. Surprisingly, the share of negative reviews dropped while positive reviews rose during COVID-19 compared to pre-pandemic times. This might be attributed to increased patient empathy for overburdened hospitals, heightened focus on infection control, and adjusted patient expectations favoring basic care delivery over non-essential aspects [66, 67].

Still, some hospital units showed disparities. The service alert unit consistently received high negative feedback, while the day surgery unit maintained higher positive ratings. This difference hints that certain services were prioritized, possibly resulting in uneven patient experiences across departments.

### Drivers of Patient Satisfaction and Dissatisfaction

Our analysis uncovered major factors that shape satisfaction or dissatisfaction. ‘Positive recognition’ consistently drove satisfaction, garnering the highest proportion of positive feedback before and during COVID-19. This finding reflects the value of recognition and gratitude in healthcare. Problems with ‘billing and accounting’, on the other hand, were a leading cause of dissatisfaction, with 68.34% negative reviews in both periods, indicating a need for clearer billing procedures and improved communication around financial matters.

We discovered that high costs for ancillary items and prescriptions contributed significantly to dissatisfaction, a phenomenon also noted in Canadian studies of out-of-pocket drug expenses [68]. Some patients also pay extra to skip long queues, intensifying their financial burden and unhappiness [68, 69]. These data call for more transparent billing practices and action on healthcare affordability.

Notably, certain themes such as ‘ICU/CCU’, ‘cardiology’, ‘radiology’, ‘families/friends’, ‘social services’, ‘discharge’, ‘information/education’, and ‘positive recognition’ saw lower positive feedback during COVID-19 compared to previous years, implying that these services struggled to sustain patient satisfaction amid pandemic-related disruptions.

### Demographic Disparities and Health Equity

Our examination of hospitals in areas with different percentages of visible minorities and low-income populations revealed clear differences in patient satisfaction patterns. From 2015 to 2020, facilities in regions with higher minority and low-income concentrations generally had better reviews, but this reversed in 2021–2022, with these same hospitals receiving considerably worse feedback. This abrupt shift may reflect the following structural imbalances:

#### Resource Gaps

Financial constraints have worsened, as several Ontario hospitals face significant challenges balancing their budgets [70], limiting opportunities for expansions or improved services. Infrastructure shortfalls further compound the crisis, with many hospital buildings in poor condition and in need of repair [71]. The pandemic hit all hospitals hard, but those serving minority and low-income areas experienced especially acute pressures. These safety-net hospitals, already underfunded, lacked the capacity to handle surging demands and were forced to halt income-generating elective services, compounding their financial strain [72].

#### Unequal Health Impacts

Studies show that visible minority groups had higher rates of COVID-19 infection, hospitalization, and mortality [73–76]. This likely fueled higher negative feedback from these communities.

#### Socioeconomic Pressures

Crowded housing, job instability, and limited preventive care made low-income communities more vulnerable to COVID-19 [77]. Hospitals in these areas faced chronic resource shortages, further stretching staff and supply chains [78, 79].

#### Language Barriers

Non-English speakers faced substantial communication challenges. One U.S. study found a two-fold increase in COVID-19 hospitalizations and over two times higher risk of death among non-English speakers [80].

#### Trust Issues

Historical mistreatment of minority communities contributed to vaccine hesitancy and reluctance to seek care. Statistics Canada found that only 56.4% of the Black population was open to vaccination, compared to 77.7% of White and 82.5% of South Asian populations [81].

These findings point to the need for resilient healthcare systems that prioritize equity, particularly in times of crisis. Future work should identify robust care models for marginalized populations and explore how social determinants of health shape patient experiences.

### Implications for Healthcare Management and Policy

#### AI-enhanced pain management ecosystems

An AI-based approach to pain management could transform patient care by using patient histories, treatment data, and real-time vitals to generate personalized pain relief strategies. This might help the 20% of Canadians who experience chronic pain and reduce the annual $38.2–$40.3 billion cost of pain management [82]. Yet, biases in AI algorithms, data privacy concerns, and the potential loss of human empathy remain real risks [83–85]. Emerging Canadian reports also convey the potential for rapid AI integration in healthcare settings could reshape patient-provider dynamics [86], while another recent study found that real-time AI alerts cut patient deaths by 43% [87].

#### Integrated mental health frameworks

Embedding mental health screening into regular medical workflows could address the uptick in ‘anxiety’ found in negative reviews from 2017 to 2022. With one in five Canadians facing mental illness each year, and around 4,000 annual suicides [88], building mental health support into hospital care could improve both physical and mental health outcomes.

#### Predictive emergency care systems

Advanced data analytics in emergency departments could improve patient flow and resource deployment. This strategy may counter the growing negative reviews in the Emergency unit, which rose from 36.1% in 2017 to 44.3% in 2021. Predictive models can forecast patient volumes with up to 90% accuracy [89], helping reduce wait times. Simulation-based methods and machine learning also show promise for identifying bottlenecks and providing real-time wait-time predictions [90, 91], potentially increasing patient satisfaction in Canadian Emergency Departments (EDs), where 90% of non-admitted patients finish their visit within 9.1 hours [92]. Interestingly, after adopting SurgeCon, a platform that uses real-time analytics to monitor capacity and predict surges, Carbonear General Hospital in Newfoundland and Labrador cut physician wait times from 104 to 42 minutes and total ED time from 199 to 134 minutes [93].

#### Blockchain-enabled billing transparency

Blockchain, a decentralized digital ledger, provides real-time updates across a peer-to-peer network with tamper-resistant data integrity [94]. This technology is particularly suited to addressing healthcare billing challenges, where clarity and trust are paramount. Blockchain-based billing systems can provide transparent, immutable records, reducing disputes and enabling patients to better understand their medical expenses. This is especially critical given the significant patient dissatisfaction (68.34%) related to billing issues. Beyond transparency, the financial benefits are substantial. Research suggests that blockchain could save the healthcare industry $20 billion annually by streamlining data management and reducing inefficiencies [95]. Real-world implementations also showcase its potential. In 2019, Anthem, the second-largest health insurer in the U.S., announced plans to use blockchain to securely manage health data for 40 million patients [96]. Similarly, the United Arab Emirates (UAE) became the first nation to integrate blockchain and AI into its organ transplant system [97], further demonstrating the transformative potential of this technology.

#### Recognition-centric care environments

Environments that value staff recognition could boost patient satisfaction, reinforcing the 73.18% positive reviews tied to positive recognition during COVID-19. Studies suggest that better staff appreciation can lead to lower turnover rates and better patient safety [98, 99]. However, the pandemic showed that recognition alone cannot solve deeper challenges like heavy workloads, stress, and inadequate pay [100, 101]. For example, 90% of nurses reported increased workload since COVID-19 started, and 57% faced financial difficulties [101]. A comprehensive strategy that addresses these root issues is essential for lasting improvements in workforce morale and patient experiences.

#### Adaptive care matrices for specialized units

Flexible care frameworks in specialized areas such as dentistry or radiology could guard against service disruptions. Hospitals that shifted resources quickly during the pandemic saw fewer bottlenecks and better patient outcomes [102–106]. Implementing these strategies may prevent spikes in negative reviews when normal operations are disrupted.

#### Health equity zone frameworks

Targeted healthcare programs in high-minority regions might reverse the decline in positive feedback during the pandemic and honor Canada’s goal to reduce health inequalities. Research indicates that indigenous and low-income groups experience higher rates of mental illness and worse health outcomes [107], calling for policy interventions like equity zones to improve care and well-being [108, 109].

### Limitations

This study offers valuable insights but has certain limitations. First, the absence of healthcare provider perspectives limits our view of the factors that shape satisfaction, although our analysis of patient reviews across sentiment, clinical entities, and themes provides a broad perspective. Second, our lack of detailed demographic data on patient ethnicity and gender might introduce bias, so we utilized regional statistics on visible minorities and low-income populations to approximate healthcare equity. Third, not every patient leaves a review, and dissatisfied individuals are often more likely to do so, which may skew findings.

Fourth, we removed incomplete data, potentially excluding some experiences, and ambiguous hospital unit names required approximate mappings, possibly introducing inaccuracies. Fifth, the dataset from 2015 is relatively small (about 500 reviews), which may limit robustness for that year, but it still revealed consistent trends. Sixth, we do not know which sites are teaching hospitals, a factor that can affect patient perceptions. Finally, without contextual information about each patient’s circumstances, we cannot fully dissect the roots of their feedback. However, we employed advanced natural language processing methods to extract deeper insights, partly offsetting this gap.

Despite these constraints, our findings establish a strong platform for future investigations. They spotlight key areas like staff interactions, billing transparency, and health equity, offering clear pathways for policy and practice improvements.

## Conclusion

This analysis of patient reviews indicates major gaps in hospital patient satisfaction before and during the COVID-19 pandemic, with many hospitals struggling to meet rising patient expectations. Staff behaviour stands out as a decisive factor, as kindness, professionalism, and helpfulness shape positive perceptions. Hospitals that invest in empathy, improved communication, and patient-focused care may see meaningful boosts in satisfaction. Meanwhile, the pandemic affected patient satisfaction unevenly across hospital units: some maintained higher positive ratings, while others encountered recurring problems linked to anxiety and infection. The unexpected dip in negative reviews, alongside a rise in positive ones, hints at greater public empathy and possible advancements in service quality, but it also illustrates how quickly patient expectations can shift during crises.

Regional disparities further complicate the picture, as satisfaction patterns switched course during the pandemic, stressing the importance of coordinated health equity measures. These findings offer a clear framework for hospitals aiming to refine patient experiences, by tightening communication, resolving shortcomings in low-performing units, encouraging staff recognition, and tackling billing challenges. Interestingly, a recent Ontario-based study found that hospitals rarely shared patient feedback with the public, limiting opportunities for improvement [40]. Relying on ongoing analysis of patient reviews, supported by NLP techniques, can identify emerging concerns early and direct adjustments that keep pace with changing community needs.

## Data Availability

The data underlying the findings of this study are available upon request. Access to the data may be granted by contacting the corresponding author.

## Acknowledgements

This work was supported by the Institute for Pandemics at the University of Toronto, the Natural Sciences and Engineering Research Council of Canada (NSERC), and a scholarship from the Vector Institute.

